# The differential impact of the COVID-19 epidemic on Medicaid expansion and non-expansion states

**DOI:** 10.1101/2021.02.23.21252296

**Authors:** Muhammad Ragaa Hussein, Islam Morsi, Engy A. Awad, Dina A. Fayed, Thamer AlSulaiman, Mohamed Habib, John R. Herbold

## Abstract

Medicaid expansion is a federally-funded program to expand health care access and coverage to economically challenged populations by increasing eligibility to Medicaid enrollment and investing in public health preventive services in the individual states. Yet, when the COVID-19 epidemic plagued the country, fourteen states were practicing their chosen decision not to enact the Medicaid expansion policy. We examined the consequences of this nationwide split in Medicaid design on the spread of the COVID-19 epidemic between the expansion and non-expansion states. Our study shows that, on average, the expansion states had 217.56 fewer confirmed COVID-19 cases per 100,000 residents than the non-expansion states [-210.41; 95%CI (−411.131) - (−2.05); P<0.05]. Also, the doubling time of COVID-19 cases in Medicaid expansion states was longer than that of non-expansion states by an average of 1.68 days [1.6826; 95%CI 0.4035-2.9617; P<0.05]. These findings suggest that proactive investment in public health preparedness was an effective protective policy measure in this crisis, unsurpassed by the benefits of COVID-19 emergency plans and funds. The study findings could be relevant to policymakers and healthcare strategists in non-expansion states considering their states’ preparations for such public health crises.

## Introduction

In April 2020, because of the COVID-19 epidemic, every state and the District of Columbia had unemployment rates that exceeded their Great Recession levels. The nation’s unemployment rate reached an unprecedented level (14.8%) since 1948 when data collection first started. The impact was more rigid on the workforce’s vulnerable sects, including teenagers, part-time workers, women, and minorities, compared to their other counterparts. A Congressional Research Service report, published in January 2021, showed that many of those unemployment gaps persisted in December 2020.^1^

The unemployment surge added to the public health crisis because employment and healthcare are twiddled together for most Americans, almost half of whom get their health insurance from employer-sponsored plans.^2^ Half of those insured postponed treatment for a family member or themselves because of cost considerations before the pandemic.^2^ As the recession escalated, many found themselves eligible for ACA coverage, increasing pressure on the state-funded Medicaid. In a phased response to the crisis, Congress passed critical pieces of legislation, including The Families First Coronavirus Response Act (FFCRA) and the Coronavirus Aid, Relief, and Economic Security (CARES) Act. The FFCRA required all insurance plans to cover COVID-19 testing without billing the beneficiaries any form of cost-sharing, such as deductibles and copayments. CARES act established the Provider Relief Fund to reimburse the participating providers for COVID-19 testing costs at Medicare rates if the patient was uninsured.^3^ Yet, it does not seem that the Congressional emergency COVID-19 funds entirely substituted the benefit of having health insurance for groups burdened with health disparities and disproportionate shares in chronic disease comorbidities, such as the African American population.^4^ In 2019, the African American population’s rate in the uninsured population was higher than the national average in ten non-expansion states. In January 2021, almost a year after Congress’s first response to the epidemic, the African American share of COVID-19 deaths was more than their share in the population in ten of the fourteen non-expansion states. ^5,6^

Most studies found that Medicaid expansion had positive associations with health care equity, access, service utilization, and self-reported health, along with reducing all-cause mortality.^7^ These improvements could be most relevant in a pandemic situation. Individuals with some form of health insurance can seek health care services early in the disease cycle, increasing their chances of positive outcomes and decreasing the number of people spreading the virus.^8^ Some state officials in expansion states credited Medicare expansion for giving their states a fighting chance against the COVID-19 epidemic that would not have been possible had their states decided not to expand their Medicaid.^9^

The pandemic’s shifting spread pattern through 2020 seemed to support the hypothesis that Medicaid expansion positioned the expansion states for better aftermath. Over summer 2020, Florida and Texas, the largest non-expansion states, were among the nation’s epicenters of the pandemic. In the fall, as the pandemic flared in the nation’s northern regions, non-expansion states of Tennessee, South Dakota, Wisconsin, and Wyoming experienced a surge in their caseloads.^10,11^ In this study, we further investigated this hypothesis and examined if there has been a differential in the COVID-19 epidemic spread between Medicaid expansion and non-expansion states.

## Study data

The unit of observation was daily U.S. state or its equivalent. Our data set tracked the 50 states and the District of Colombia over three hundred and seven days, starting January 22, 2020, and ending November 23, 2020. The predictor variable was a state’s implementation of the Medicaid expansion. Thirty-three states, and the District of Columbia, implemented the Medicaid expansion, while 14 states did not. We included Oklahoma and Missouri states in the non-expansion category because they have not implemented, though approved, expanding their Medicaid by the Time the COVID-19 epidemic hit. Voters in the State of Oklahoma approved an amendment to the state’s constitution on June 30, 2020, to expand Medicaid starting no later than July 1, 2021; and voters in the State of Missouri approved a ballot measure to the same effect yet starting no later than March 1, 2021.^12^

### Outcomes of interest

Our first outcome of primary interest was the daily count of cases per 100,000 residents. We collected this information from the COVID-19 resource center provided by Johns Hopkins Center for Systems Science and Engineering. Our second outcome of interest was the doubling time of the COVID-19 case count. The doubling time is the length of the period it takes for confirmed cases to double in number. It is a crucial indicator epidemiologists use to assess how rapidly an epidemic could spread in the population. This measure also indicates the urgency of the situation and the magnitude of public health efforts needed to control the epidemic situation.^13^

### Covariates

We added information on the at-risk adults as a share of all adults ages 18 and older. We preferred to use this measure rather than the total population size in light of the literature’s current consensus that not all age groups are equally vulnerable to COVID-19^14, 15, 16, 17^. We included the number of community health centers’ delivery sites as a proxy of the state’s primary care capacity to the underserved populations.^17^ We also had information on the length of active stay-home orders as a proxy of the respective state’s epidemic containment policies’ stringency. Finally, we controlled for the proportion of surveyed population always wearing masks. We obtained this data from the New York Times survey in July 2020.^18,19^

## Methods

We applied a multi-step model to investigate the research hypothesis. First, we used a Generalized Estimating Equation model to examine the population-averaged relationship between the cumulative count of daily COVID-19 cases per 100,000 state residents and Medicaid expansion (predictor variable) in the different states. Second, we calculated the COVID-19 case doubling time for each state through the study period. (Table 1) We used the COVID-19 case doubling time as the response variable in a Generalized Estimating Equation model to examine its relationship with the respective states’ enactment of Medicaid expansion. We used SAS 9.4 TS 1M6 by SAS Institute to conduct these analyses and visualize the findings.

**Table 1:** COVID-19 cases Doubling Time through the study period, by state and Medicaid Expansion status

| Medicaid Expansion | State | Doubling Time |
| --- | --- | --- |
| Enacted | Alaska | 20.9 |
|  | Arizona | 13.3 |
|  | Arkansas | 19.2 |
|  | California | 14.2 |
|  | Colorado | 21.8 |
|  | Connecticut | 23.0 |
|  | Delaware | 21.5 |
|  | District of Columbia | 25.9 |
|  | Hawaii | 21.2 |
|  | Idaho | 20.0 |
|  | Illinois | 14.0 |
|  | Indiana | 19.2 |
|  | Iowa | 18.8 |
|  | Kentucky | 18.6 |
|  | Louisiana | 23.1 |
|  | Maine | 25.4 |
|  | Maryland | 19.2 |
|  | Massachusetts | 15.5 |
|  | Michigan | 23.9 |
|  | Minnesota | 17.7 |
|  | Montana | 21.2 |
|  | Nebraska | 17.9 |
|  | Nevada | 18.7 |
|  | New Hampshire | 21.2 |
|  | New Jersey | 21.5 |
|  | New Mexico | 20.6 |
|  | New York | 22.8 |
|  | North Dakota | 19.3 |
|  | Ohio | 20.2 |
|  | Oregon | 19.4 |
|  | Pennsylvania | 20.5 |
|  | Rhode Island | 19.3 |
|  | Utah | 18.7 |
|  | Vermont | 27.4 |
|  | Virginia | 19.0 |
|  | Washington | 16.0 |
|  | West Virginia | 22.1 |
| Not<br>Enacted | Alabama | 18.9 |
|  | Florida | 16.5 |
|  | Georgia | 17.9 |
|  | Kansas | 18.5 |
|  | Mississippi | 19.7 |
|  | Missouri | 18.6 |
|  | North Carolina | 16.7 |
|  | Oklahoma | 17.8 |
|  | South Carolina | 17.8 |
|  | South Dakota | 20.6 |
|  | Tennessee | 17.9 |
|  | Texas | 16.7 |
|  | Wisconsin | 20.0 |
|  | Wyoming | 22.3 |

Source: Authors' analysis of US COVID-19 data, obtained from the COVID-19 resource center provided by Johns Hopkins Center for Systems Science and Engineering.

## Results

### Descriptive statistics

The public health layout differed in the non-expansion states from that in the expansion states. On average, the non-expansion states had a relatively higher ratio of the hospital and ICU beds to their population than the Medicaid expansion states. Yet, the non-expansion states also had a larger uninsured population and fewer primary care Community Health Centers delivery sites than the expansion states. Compared to the expansion states, the non-expansion states mandated stay-home orders for a shorter average period. Also, non-expansion states’ residents’ compliance to wearing masks was not as substantial as residents in the expansion states. (Table 2) The demographic profile of the two groups of states was comparable but slightly different. In 2019, non-white racial minorities in non-expansion states made 33.6% of the population, compared to 32% in expansion states. The largest minority in non-expansion states was African Americans, 15%, followed by Hispanics, 11%; while the largest minority in expansion states was Hispanics, 12.6%, followed by African Americans, 9.2%.

**Table 2:** Indicators of the public health profiles in Medicaid expansion and non-expansion states by 2019 averages.

|  | Nonexpansion states | Expansion states |
| --- | --- | --- |
| Uninsured rate in the population | 11.6% | 7.2% |
| Hospital Beds per 10,000 Population | 28.7 | 24.1 |
| ICU Beds per 10,000 Population | 2.9 | 2.6 |
| Community Health Centers Delivery Sites | 222.8 | 229.4 |
| Stay-home duration | 26.1 | 76.5 |
| Proportion of population always wearing masks | 46.8% | 58.5% |

Source: Authors' analysis of study data sourced from multiple Kaiser Family Foundation reports on 2019 state data.

### Inferential statistics

The study models’ results confirm, with statistical significance, that Medicaid expansion states had a lower caseload of COVID-19 cases than the non-expansion states by an average of 210.4 cases per 100,000 residents. [-210.41; 95%CI (−411.131) - (−2.05); P<0.05]. (Figure 1) Also, it took 1.68 additional days, on average, for the COVID-19 cases in Medicaid expansion states to double in number, compared to the non-expansion states [1.6826; 95%CI 0.4035-2.9617; P<0.05]. (Figure 2)

**Figure 1:**
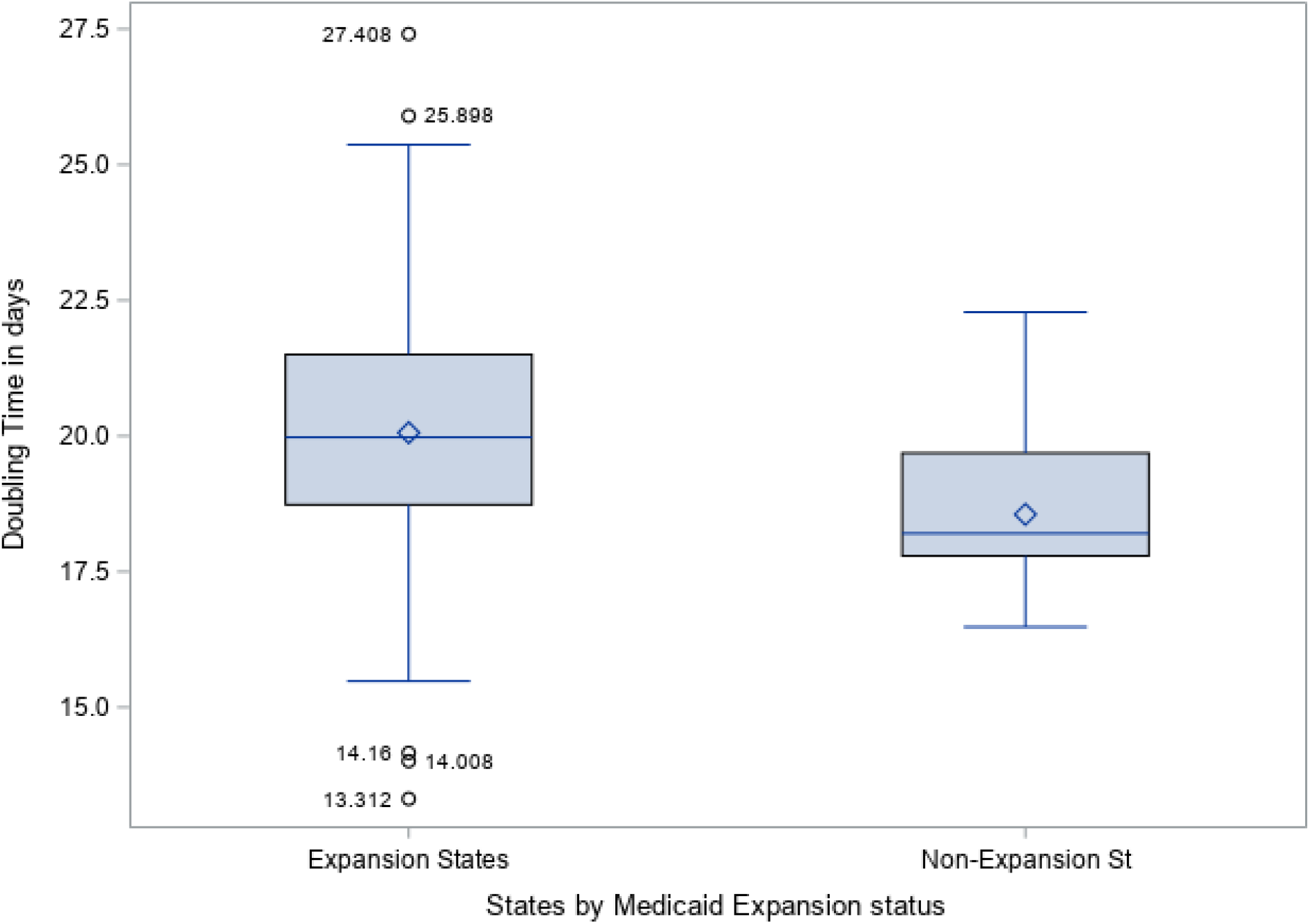
Average COVID-19 cases Doubling Time in Medicaid Expansion and the Non-Expansion States through the study period Source: Authors’ analysis of study data, sourced from the COVID-19 resource center provided by Johns Hopkins Center for Systems Science and Engineering.

**Figure 2:**
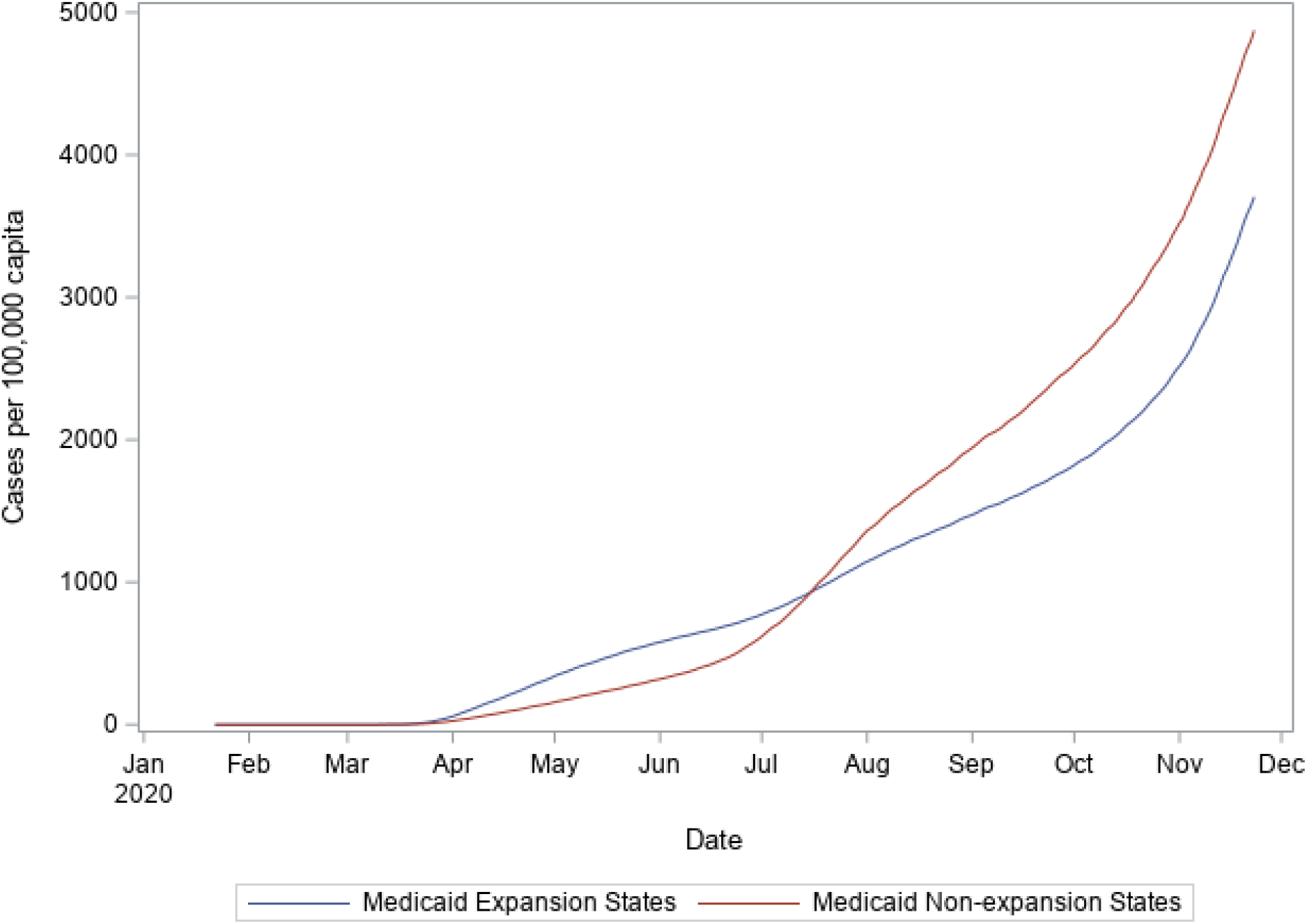
COVID-19 Cumulative Caseload over Study Time, by States Medicaid Expansion Source: Authors’ analysis of study data, sourced from the COVID-19 resource center provided by Johns Hopkins Center for Systems Science and Engineering.

## Conclusion

The findings present an opportunity for the U.S. to improve its health care system, building on the ongoing efforts to ensure accuracy of its COVID-19 counting; an advantage not available in many other countries.^20^ The results show that COVID-19 epidemic spread at a significantly slower rate in the expansion states than the non-expansion states. It took almost two additional days for COVID-19 case numbers to double in Medicaid expansion states than in the non-expansion states. The epidemic also caught a considerably lower number of confirmed cases in expansion states than in the non-expansion states, by about 210 confirmed COVID-19 cases per 100,000 state residents (Figure 2). The two groups of states differed in their public health infrastructure and their COVID-19 containment policies’ stringency. The data analysis suggests that the expansion states, on average, had a slightly bigger primary care provider capacity and significantly lower uninsured rates than the non-expansion states, which had bigger secondary care capacity than their counterparts. The average duration of stay-home orders in the expansion states was threefold as in the non-expansion states, whose residents seemed less compliant to wearing masks.

It’s plausible that the observed advantage of Medicaid expansion states could be stemming from multiple layers in the U.S. health care system. Besides the differences in primary care capacity and uninsured rates, the multistep Congressional response does not seem to have entirely replaced the benefit of extended Medicaid health insurance coverage for residents in non-expansion states. The CARES Act reimbursed the providers for treating uninsured COVID-19 patients, and the law conditioned eligibility for having a primary diagnosis of COVID-19. The reimbursement was also conditioned on the availability of funds, not granted, and providers’ enrollment was made optional. The limitation these conditions might have posed on the uninsured population’s access to care in the epidemic probably weighed most on the states with the highest uninsured rates, the non-expansion states. (Table 2)

Also, not everyone losing their job in non-expansion states would qualify for Medicaid. Many probably fall into the “coverage gap”; a void space in ACA coverage entrapping people of income below the federal poverty level but higher than Medicaid requirements. This sect of the economically challenged population is ineligible for both Medicaid and subsidized coverage in the marketplace. In 2019, most of the uninsured in non-expansion states fell into the coverage gap, which only exists because their states have opted not to expand their Medicaid. People with health insurance could seek medical assistance and diagnosis should they have early COVID-19 symptoms. The early diagnosis of confirmed COVID-19 cases could allow them to quarantine themselves early in the disease cycle, potentially limiting their role in spreading the virus.

On the other hand, the positive COVID-19 cases in the uninsured population could unknowingly spread the virus until they had a confirmed COVID-19 diagnosis to become eligible for federal assistance. The confirmed COVID-19 diagnosis would typically follow a period of asymptomatic, then symptomatic, spread of the virus. This could explain why states with the highest uninsured rates, the non-expansion states, had a faster spread of the epidemic in their residents, as the results show. (Table 1) This risk might have been at its highest early in the epidemic when testing was not widely available; and in the months before the previous administration deployed the provider reimbursement program in April 2020.

State officials in many of the expansion states had a window of years to work on fortifying their states’ public health reach for several years, between when the Medicaid expansion became effective, in 2014, and when the epidemic hit in late 2019. This period could have given the public health workers in those states enough time to extend their reach to the vulnerable population of known health disparities and for the beneficiaries to learn of their options and connect with their health care providers. This luxury of time and reach was not equally available to the economically challenged people in the non-expansion states. The federal assistance they received came after the epidemic has already struck the country.

Finally, this study’s results bring the limelight to the high cost of fragmentation in the U.S. healthcare delivery system at the national level. States with a common ground of health policy features collectively outperformed the others regarding COVID-19 rate of spread and caseload. This gap in COVID-19 toll will probably translate to a widening of the existent gap between the two groups in health outcomes in favor of the expansion states, especially with the spike in demand for mental health services, which Medicaid expansion already covers. In a recent survey by Kaiser Family Foundation, conducted in July 2020, 53% of U.S. adult respondents reported a negative impact on their mental health from epidemic-related stress conditions like isolation and job loss worries. The reported figure in March 2020 was 32%. Medicaid expansion already covers those services, putting the expansion states in an advantageous position in the epidemic’s aftermath. Medicaid patients and the uninsured in non-expansion states will not have the same support as their counterparts in expansion states.

It’s noteworthy that this fragmentation is not the only one the health care system has. Another fragmentation in the payer system already caused the U.S. to have the most expensive health care service in the world, per most of the literature and multiple OECD economic reports. This repeatedly proven need for a more streamlined and less fragmented system could be significant to the current administration for the political consideration of introducing instating public insurance options, or a universal health care choice, in the U.S. health care market.

In conclusion, this study confirms that Medicaid expansion states were in a better public health position for crises like the COVID-19 epidemic. While after-the-fact federal assistance could have helped many uninsured, its benefit was not a full replacement to public health preparedness in terms of expanded insurance coverage. Further scientific inquiry will be needed to investigate further the Medicaid expansion elements that most helped the expansion states collectively outperform their non-expansion counterparts.

## Limitations

This study has several limitations. First, widespread COVID-19 screening was not evenly available through the study period. It is possible that the case records early in the epidemic are not entirely indicative of COVID-19 spread. Second, we included some, but not all, of the epidemic containment state policies, and we did not add a measure of the stringency of enforcement of these local measures in the respective states. Third, we included one measure of primary care capacity, the number of community health centers’ delivery sites. Still, there are other capacity elements like the number of primary care offices and registered general practitioners. Fourth, the analyses did not include the states’ financial standings that could play into a state’s decision of embracing Medicaid Expansion or not. Finally, the study is descriptive and did not divulge into the causality underlying the findings.

## Data Availability

The data will be made available on the author team's website www.researchocrats.com/data for use with proper citation. Till then, we'll gladly share the data upon request.

## References

1. Falk G, Carter J, Nicchitta I, Nyhof E, Romero P. Unemployment Rates During the COVID-19 Pandemic: In Brief. Congr Res Serv. 2020:2–16. https://crsreports.congress.gov.

2. Hamel L, Muñana C, Brodie M. Kaiser Family Foundation / LA Times Survey Of Adults With Employer-Sponsored Health Insurance. 2019;(May).

3. King JS. Covid-19 and the Need for Health Care Reform. N Engl J Med. 2020;382(26):e104. doi:10.1056/NEJMp2000821

4. Thorpe KE, Ko Chin K, Cruz Y, Innocent MA, Singh L. The United States can reduce socioeconomic disparities by focusing on chronic diseases. Heal Aff Blog https://wwwHealorg/do/101377/hblog20170817. 2019;61561.

5. The Atlantic. The COVID Tracking Project. https://covidtracking.com/race.

6. Schwartz K, Tolbert J. Limitations of the Program for Uninsured COVID-19 Patients Raise Concerns.; 2020. https://www.kff.org/coronavirus-policy-watch/limitations-of-the-program-for-uninsured-covid-19-patients-raise-concerns/#.

7. Artiga S, Damico A, Garfield R. The Impact of the Coverage Gap for Adults in States Not Expanding Medicaid by Race and Ethnicity.; 2015. http://files.kff.org/attachment/issue-brief-the-impact-of-the-coverage-gap-for-adults-in-states-not-expanding-medicaid-by-race-and-ethnicity.

8. Chakrabarti R, Meyerson L, Nober W, Pinkovskiy M. The Affordable Care Act and the COVID-19 Pandemic: A Regression Discontinuity Analysis. SSRN Electron J. 2020;(948). doi:10.2139/ssrn.3733129

9. Gee R. Aligning Public Health Infrastructure and Medicaid to Fight COVID-19. Am J Public Health. 2020;110(S2):S173. doi:10.2105/AJPH.2020.305826

10. Brown C. Medicaid Expansion Ballot Measures Brewing in Three More States. Bloomberg Law. https://news.bloomberglaw.com/health-law-and-business/medicaid-expansion-ballot-measures-brewing-in-three-more-states. xPublished 2021.

11. Center for America Progress. The Pandemic and Economic Crisis Are Wake-Up Call for State Medicaid Expansion - Center for American Progress. https://www.americanprogress.org/issues/healthcare/news/2020/11/09/492808/pandemic-economic-crisis-wake-call-state-medicaid-expansion/. xPublished 2021. accessed January 31, 2021.

12. Kaiser Foundation. Status of State Action on the Medicaid Expansion Decision. https://www.kff.org/medicaid/issue-brief/status-of-state-medicaid-expansion-decisions-interactive-map/#:~:text=CoverageundertheMedicaidexpansion,%2CVirginia(1%2F1%2F.

13. Galvani AP, Lei X, Jewell NP. Severe acute respiratory syndrome: Temporal stability and geographic variation in case-fatality rates and doubling times. Emerg Infect Dis. 2003;9(8):991–994. doi:10.3201/eid0908.030334

14. Centers for Disease Control and Prevention (CDC). Certain Medical Conditions and Risk for Severe COVID-19 Illness. https://www.cdc.gov/coronavirus/2019-ncov/need-extra-precautions/people-with-medical-conditions.html. xPublished 2020. accessed January 31, 2021.

15. The Centers for Medicare & Medicaid Services (CMS). COVID-19 Resources on Vulnerable Populations for Consumers and Patients. https://www.cms.gov/About-CMS/Agency-Information/OMH/resource-center/COVID-19-Resources/COVID-19-Resources-For-Consumers-And-Patients. xPublished 2020. accessed January 31, 2021.

16. WHO Western Pacific. COVID-19 advice - High risk groups. https://www.who.int/westernpacific/emergencies/covid-19/information/high-risk-groups. xPublished 2020. accessed January 31, 2021.

17. Kaiser Foundation. State COVID-19 Data and Policy Actions. https://www.kff.org/coronavirus-covid-19/issue-brief/state-covid-19-data-and-policy-actions/. xPublished 2021. accessed January 31, 2021.

18. Josh Katz MS-K and KQ. A Detailed Map of Who Is Wearing Masks in the U.S. The New York Times. https://www.nytimes.com/interactive/2020/07/17/upshot/coronavirus-face-mask-map.html. xPublished 2020. accessed January 31, 2021.

19. The New York Times. Mask-Wearing Survey Data. The New York Times. https://github.com/nytimes/covid-19-data/tree/master/mask-use. xPublished 2020. accessed January 31, 2021.

20. Hussein MR, Mba M, Alsulaiman T, et al. The Relationship between Democracy embracement and COVID-19 reported casualties worldwide. doi:10.1101/2021.01.11.21249549

